# Differentially expressed circular RNAs in peripheral blood mononuclear cells of PD patients

**DOI:** 10.1101/2020.11.14.20231779

**Authors:** Stylianos Ravanidis, Anastasia Bougea, Dimitra Karampatsi, Nikolaos Papagiannakis, Matina Maniati, Leonidas Stefanis, Epaminondas Doxakis

**Author notes:** Corresponding author: Epaminondas Doxakis, Corresponding author’s address: Center of Basic Research, Biomedical Research Foundation, Academy of Athens, Soranou Efesiou 4, Athens, 11527, Greece., Corresponding author’s, Corresponding author’s.

## Abstract

**Background:** New noninvasive and affordable molecular approaches that will complement current practices and increase the accuracy of PD diagnosis are urgently needed. CircRNAs are highly stable non-coding RNAs that accumulate with aging in neurons and are increasingly shown to regulate all aspects of neuronal development and function.

**Objectives:** The aims of the present study were to identify differentially expressed circRNAs in PBMCs of idiopathic PD patients and explore the competing endogenous RNA networks affected.

**Methods:** Eighty-seven circRNAs were initially selected based on relatively high gene expression in the human brain. Over half of these were readily detectable in PBMCs using RT-qPCR. Comparative expression analysis was then performed in PBMCs from sixty controls and sixty idiopathic subjects with PD.

**Results:** Six circRNAs derived from MAPK9, HOMER1, SLAIN1, DOP1B, REPS1, and PSEN1 transcripts were significantly downregulated in PD patients. The classifier that best distinguished PD consisted of four circRNAs with an AUC of 0.84. CLIP-Seq data revealed that the RNA binding proteins bound by most of the deregulated circRNAs include the neurodegeneration-associated FUS, TDP43, FMR1 and ATXN2. MicroRNAs predicted to be sequestered by most deregulated circRNAs had the GOslim categories ‘Protein modification’, ‘Transcription factor activity’ and ‘Cytoskeletal protein binding’ mostly enriched.

**Conclusions:** This is the first study that identifies circRNAs deregulated in the peripheral blood of PD patients. They may serve as diagnostic biomarkers and since they are highly expressed in the brain and are derived from genes with essential brain functions, they may also hint on the PD pathways affected.

## INTRODUCTION

The diagnosis of PD is currently based on clinical diagnostic criteria and neuroimaging and is monitored by rating scales related to motor and non-motor features ^1^. Rating scales are frequently subjective and influenced by periodic fluctuations in symptoms and effective symptomatic therapies, while neuroimaging techniques, such as DAT-SPECT, offer a quantifiable measure of disease progression but are limited by practicality and costs ^2^. In addition, protein biomarkers, such as those based on alpha-synuclein (SNCA) and dopamine metabolic products have yielded mixed results, do not reflect disease progression and require an invasive lumbar puncture ^3^.

Circular RNAs (circRNAs) are a newly recognized class of single-stranded regulatory RNAs that are formed by head-to-tail splicing in which a downstream 5’ splice site is covalently connected to an upstream 3’ splice site of an RNA molecule. The result is an enclosed non-polyadenylated circular transcript ^4-6^. Due to the lack of free ends, that are normally targeted by 3’ and 5’ exoribonucleases, circRNAs are extremely stable with half-life of more than 48 h compared to approximately 6 h of linear transcripts ^7, 8^. There are different subtypes of circRNAs including exonic, intronic and exo-intronic. Exonic circRNAs are mostly localized in the cytoplasm where they act as sponges for microRNAs (miRNAs) and RNA-binding proteins (RBPs), thus inhibiting their interaction with mRNA targets ^4, 9-11^. In contrast, intronic or exo-intronic circRNAs are mostly localized in the nucleus and have few or no binding sites for miRNAs; instead they function to control transcription ^12, 13^. Interestingly, the co-transcriptional biogenesis of circRNAs has also been shown to reduce linear host mRNA levels and change downstream splice-site choice in some mRNAs ^11, 14, 15^.

CircRNAs are widely conserved and more abundant in the brain than in any other tissue ^16^ with many being expressed in an organ-specific manner, along with their host genes which are enriched with tissue-specific biological functions ^17^. For instance, brain circRNA host genes are enriched in neurotransmitter secretion, synaptic activities and neuron maturation ^17^. Importantly, however, they are regulated independently from their linear counterparts ^16, 18^ with 60% of CNS circRNAs being upregulated throughout development, especially during synaptogenesis, while only 2% of their linear isoforms show this tendency ^17^.

Recent studies revealed the deregulation of circRNAs in neurodegenerative diseases and neuropsychiatric disorders (reviewed in ^19^). Furthermore, several brain-enriched circRNAs have been associated with pathogenetic processes of neurodegeneration. For instance, CDR1as (ciRS-7), a highly abundant circRNA in the brain, is downregulated in the brain of patients with AD ^20^. This circRNA contains 63 binding sites for miR-7 and therefore it is acting as an efficient sponge for it ^4^. Importantly, critical proteins for the neurodegeneration processes such as the ubiquitin protein ligase A (UBE2A), which catalyses the proteolytic clearing of toxic amyloid peptides in AD, and SNCA, which accumulates in PD/AD, are both targets of miR-7 ^21, 22^. More recently, another circRNA, circSLC8A1, was found to increase in the substantia nigra of individuals with PD and in cultured cells exposed to the oxidative stress-inducing agent paraquat ^23^. Importantly, circSLC8A1 carries seven binding sites for miR-128, an abundant and brain-restricted miRNA that governs neuronal excitability and motor behavior ^24-27^.

Peripheral blood mononuclear cells (PBMCs) inherit the same genetic information as brain cells and are armed with abundant signaling pathways that respond to pathological changes. Multiple studies have shown that genome-wide transcriptional and alternative splicing profiles in peripheral blood parallel changes in gene expression in the brain, reflecting broad molecular and cellular impairments ^28-33^. Therefore, PBMCs provide a powerful and minimally-invasive tool for the identification of novel targets for neurodegeneration research. Considering that circRNAs a) are abundant in the brain modulating gene expression *en masse*, b) are stable, c) do not get modified like proteins, hence levels directly correlate with activity and d) can be accurately quantified by routine and fast laboratory methods, such as RT-qPCR, suggests that they not only represent excellent candidate biomarkers but also important constituents of the pathophysiological processes implicated in neurological diseases. The purpose of this study was to identify differentially expressed brain-enriched circRNAs in PBMCs from patients with idiopathic PD (iPD) and pinpoint competing endogenous (ceRNA) networks.

## MATERIAL AND METHODS

Figure 1 provides a schematic representation of the workflow.

**Figure 1.**
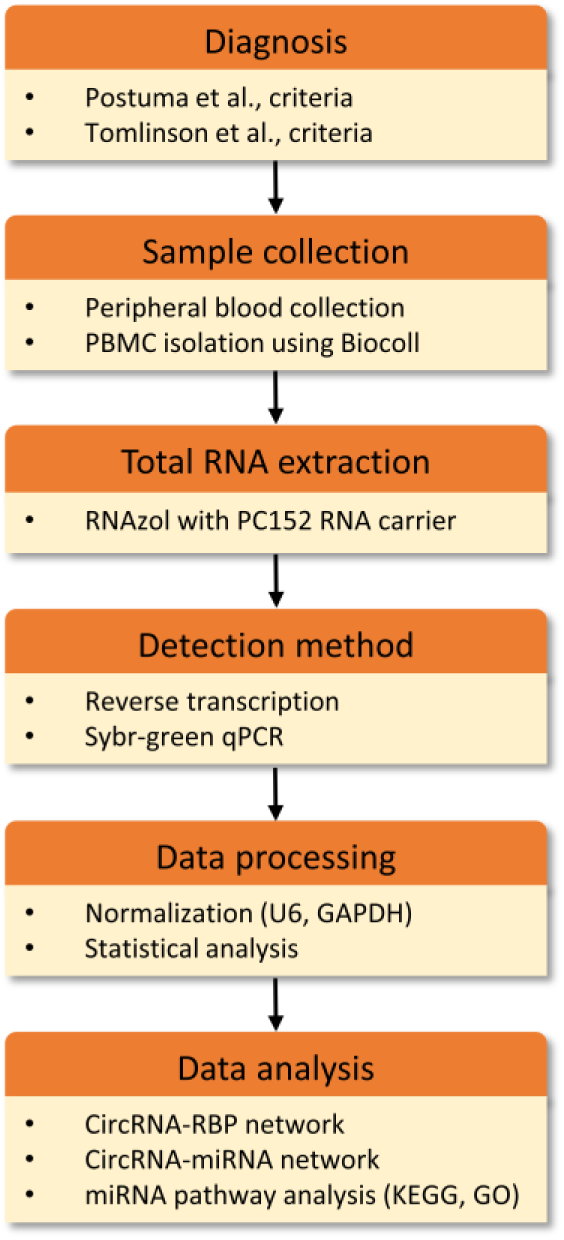
Schematic representation of the workflow.

### Study population

The present study included 60 iPD patients and 60 healthy individuals in two separate cohorts. Patients were assessed with brain magnetic resonance imaging (MRI) or computed tomography (CT) and no relevant brain vascular lesions explaining the clinical phenotype were detected. The control group included spouses or unrelated companions of patients who had no known neurological disease, comorbidities or PD family history. Individuals with concurrent malignant tumours, psychiatric disorders, collagen diseases, endocrine and cardiovascular diseases, or infections were excluded from this study, since these conditions are expected to alter the expression profile of transcripts. Patients affected by atypical parkinsonism were also excluded. All patients and controls were recruited from the National and Kapodistrian University of Athens’ First Department of Neurology at Eginition hospital. PD was diagnosed by two neurologists according to Postuma et al., criteria ^1^. In all cases, essential demographic and clinical information, including the study questionnaire for motor and non-motor manifestations of the disease, rating scales [Hoehn & Yahr (H&Y) stage, mini-mental state examination (MMSE, cognitive impairment score <26 ^34^), Unified Parkinson’s Disease Rating Scale part III (UPDRS III) in the on or off state] were collected and documented. The demographic and clinical features of patients and controls are summarized in Table 1. Levodopa equivalent daily dose (LEDD) was calculated for the patient group according to Tomlinson et al., criteria ^35^. The Eginition hospital and BRFAA ethics committees approved the study and all participants provided written consent.

**Table 1.**
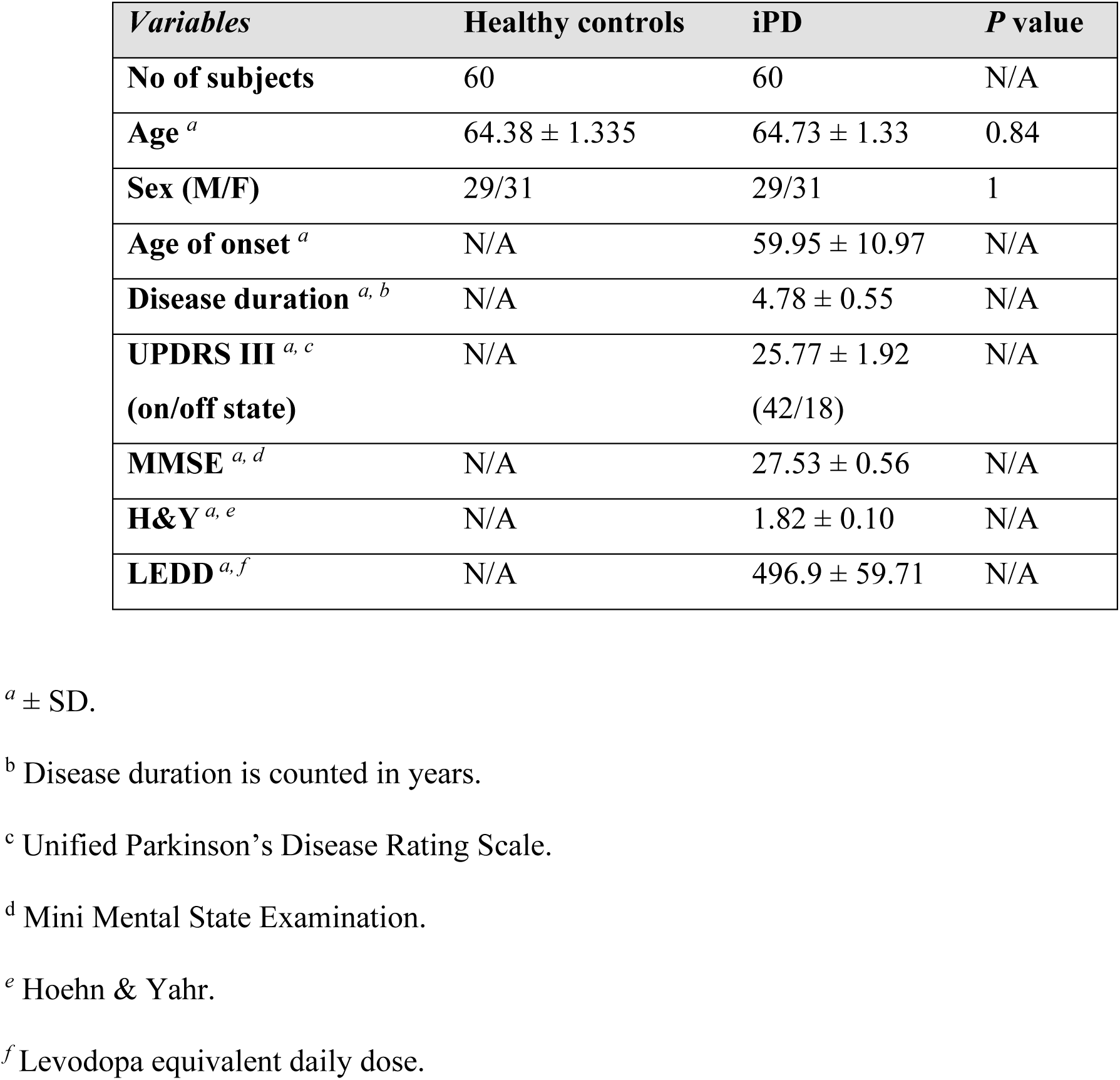
Demographic and clinical profiles of healthy controls and PD patients.

| *Variables* | **Healthy controls** | **iPD** | ***P* value** |
| --- | --- | --- | --- |
| **No of subjects** | 60 | 60 | N/A |
| **Age**<sup>a</sup> | 64.38 ± 1.335 | 64.73 ± 1.33 | 0.84 |
| **Sex (M/F)** | 29/31 | 29/31 | 1 |
| **Age of onset**<sup>a</sup> | N/A | 59.95 ± 10.97 | N/A |
| **Disease duration**<sup>a, b</sup> | N/A | 4.78 ± 0.55 | N/A |
| **UPDRS III**<sup>a, c</sup><br>(on/off state) | N/A | 25.77 ± 1.92<br>(42/18) | N/A |
| **MMSE**<sup>a, d</sup> | N/A | 27.53 ± 0.56 | N/A |
| **H&Y**<sup>a, e</sup> | N/A | 1.82 ± 0.10 | N/A |
| **LEDD**<sup>a, f</sup> | N/A | 496.9 ± 59.71 | N/A |

<sup>a</sup> ± SD.
<sup>b</sup> Disease duration is counted in years.
<sup>c</sup> Unified Parkinson's Disease Rating Scale.
<sup>d</sup> Mini Mental State Examination.
<sup>e</sup> Hoehn & Yahr.
<sup>f</sup> Levodopa equivalent daily dose.

### Isolation of peripheral blood mononuclear cells

PBMCs were isolated from whole blood by employing density-gradient centrifugation using the Biocoll Separating Solution according to manufacturer’s instructions (Biochrom AG).

### Total RNA extraction and RT-qPCR analysis

Total RNA extraction was performed using the RNAzol^®^-RT reagent according to manufacturer’s instructions (Molecular Research Center, MRC). To improve the yield of the small RNA fraction, a polyacryl carrier (PC152, MRC) was added during the extraction method. Reverse transcription reactions were performed in triplicate for every sample. Similarly, qPCR was performed in triplicate on the Roche Lightcycler^®^ 96 using the SYBR FAST Universal 2X qPCR Master Mix from Kapa Biosystems. For the differential expression analysis, we selected only those circRNAs that were detected in PBMCs with a crossing threshold (Ct) value below 30, for improved detection accuracy. All primers span the splice junction. *U6* and *GAPDH* were used as reference genes. The relative expression level of circRNAs was calculated using the 2^-ΔΔCt^ method between age- and gender-matched counterparts. Primer sequences can be found in Suppl. Table 1.

### CircRNA selection process

Eighty-seven circRNAs were carefully selected by cross-examining the data from three genome-wide surveys ^4, 18, 36^. We chose circRNAs that had high expression in the brain (Rybak-Wolf score above ∼1,000) and low or no expression in other tissues. The host gene expression was also taken into account in the selection process. Initially, based on GTEx portal expression data, circRNAs for which host transcripts were specifically expressed in the brain were selected. However, we found that many circRNAs derived from these transcripts were not readily detectable in PBMCs. We, therefore, widened the analysis to host transcripts that are brain- or at least cerebellum-enriched (i.e. not exclusively expressed in the brain). Last, we included six brain-abundant circRNAs deriving from host transcripts with low expression in the brain (UBXN7_circ_0001380, TMEM138_circ_0002058, ZNF292_circ_0004058, HAT1_circ_0008032, ZFAND6_circ_0000643, UIMC1_circ_0001558) and eight circRNAs that are hosted by brain-relevant transcripts that have been found deregulated in AD (CORO1C_circ_0000437, WDR78_circ_0006677, PHC3_circ_0001359, SLAIN2_circ_0126525) ^37^ and autism (FAM120A_circ_0001875, CSNK1G3_circ_0001522, VMP1_circ_0006508, SMARCA5_circ_0001445) ^38^. For the list of circRNAs analyzed see Suppl. Tables 1 and 2.

### CircRNA target network

The interactions of the differentially expressed circRNAs with miRNAs and RBPs were identified by obtaining data from the Circular RNA Interactome (CircInteractome) and Interactional Database of Cancer-Specific CircRNAs (IDCSC) databases, respectively ^39, 40^. CircInteractome utilizes the TargetScan algorithm to predict microRNA response elements (MREs, ie miRNA-binding sites) while IDCSC hosts circRNA cross-linking immunoprecipitation (CLIP)-Seq data for the different RBPs extracted from StarBase database ^41^. The circRNA-miRNA and circRNA-RBP interactomes were then manually curated using the Cytoscape v.3.8.0 platform.

### miRNA pathway analysis

The DIANA mirPath v.3 software suite was used to identify miRNA-regulated pathways. This software renders possible the functional annotation of miRNAs using standard hypergeometric distributions, unbiased empirical distributions and meta-analysis statistics ^42^. Here, predicted targets from DIANA microT-CDS algorithm with high quality experimentally supported interactions were used to identify KEGG molecular pathways, as well as GO terms targeted by each miRNA. The combinatorial effect of deregulated miRNAs was identified by simultaneously selecting multiple miRNAs in the software. The default values (p-value threshold 0.05, microT-CDS threshold 0.8, FDR correction option ticked) were used for the analysis.

### Statistical analysis

Statistical analysis was performed using GraphPad PRISM v5.0 and R v3.5.3. All data underwent a normality test (Shapiro–Wilk), and were found to be non-normally distributed. As a result, all circRNA data underwent a logarithmic transformation (with base 2), in order to better approximate the normal distribution. The parametric t-test was used to observe differences between healthy controls and PD patients. We applied the Benjamini – Hochberg false discovery rate correction in the resulting p-values to account for the multiple number of tests. Spearman’s method with Bonferroni correction for multiple comparisons was used to correlate circRNA expression levels with participants’ demographic and clinical characteristics (p-value threshold 0.0005).

To assess the possibility that sex is a confounding factor, the two-way ANOVA model was applied to the log-transformed data (normally distributed) with sex as an additional factor. No difference in the circRNAs that were statistically significant was found.

Receiver operating characteristic (ROC) curves were constructed and the area under the curve (AUC) was calculated to evaluate the predictive sensitivity and specificity of PBMC circRNAs for PD diagnosis. The cutoff value for the ROC analysis was determined using the Youden Index. Data are presented as means ± SEM. circRNA selection was based on the stepwise removal approach. A logistic regression statistical model containing all available circRNAs as independent variables and PD status as the dependent variable was built. Then, the circRNAs with the least contribution in the model (as determined by an F test) were removed. This process continued until no further removals were possible.

### Data availability

The datasets analysed during the current study are all available from the corresponding author on request.

## RESULTS

### circRNAs are differentially expressed in PBMCs of idiopathic PD patients

The demographic and clinical characteristics of 60 healthy controls and 60 iPD patients are summarized in Table 1. The mean age of 64.5 and the sex ratio were the same for both groups. The disease duration for the PD group was 4.8±0.55 years and the MMSE score 27.5±0.56. Initially, RT-qPCR was employed to detect plasma levels of 32 circRNAs that are highly-expressed by brain cells. It was anticipated that a sufficient quantity of brain-derived circRNAs would find its way into the plasma. However, only two circRNAs, RMST (at Ct 29) and PSD3 (at Ct 28), were detected. Using the same amount of RNA, this time extracted from human brain tissue, it was revealed that all circRNAs were readily detectable with an average Ct value of 26, demonstrating that all primer pairs were functional (data not shown). This indicated that brain-enriched circRNAs are not as abundant as brain-enriched miRNAs (average Ct value of 17.5 for 21 brain-enriched miRNAs in the same human brain total RNA) and are not circulating in appreciable amount in the blood (average Ct value for the corresponding miRNAs in the plasma is 25) ^43^.

Based on previous studies showing that genome-wide transcriptional and alternative splicing profiles in peripheral blood cells parallel changes in gene expression in the brain, the levels of circRNAs were next assessed in PBMCs. We increased the number of primer sets to 87 and found that 48 were detected with a Ct value below 30, safeguarding accurate and reproducible detection. These circRNAs were then analyzed for differential expression in healthy control and iPD patient samples (Suppl. Table 3).

Following multiple comparison adjustment, six circRNAs were significantly altered in the PBMCs obtained from PD patients compared to healthy controls. MAPK9_circ_0001566, HOMER1_circ_0006916, SLAIN1_circ_0000497, DOP1B_circ_0001187, RESP1_circ_0004368, and PSEN1_circ_0003848 were all downregulated in the PD cohort (Fig. 2 and Suppl. Table 3). The swarm plots for the 42 circRNAs whose relative expression was not significantly altered in the PBMCs of idiopathic PD patients are shown in Supplemental Fig 1.

**Figure 2.**
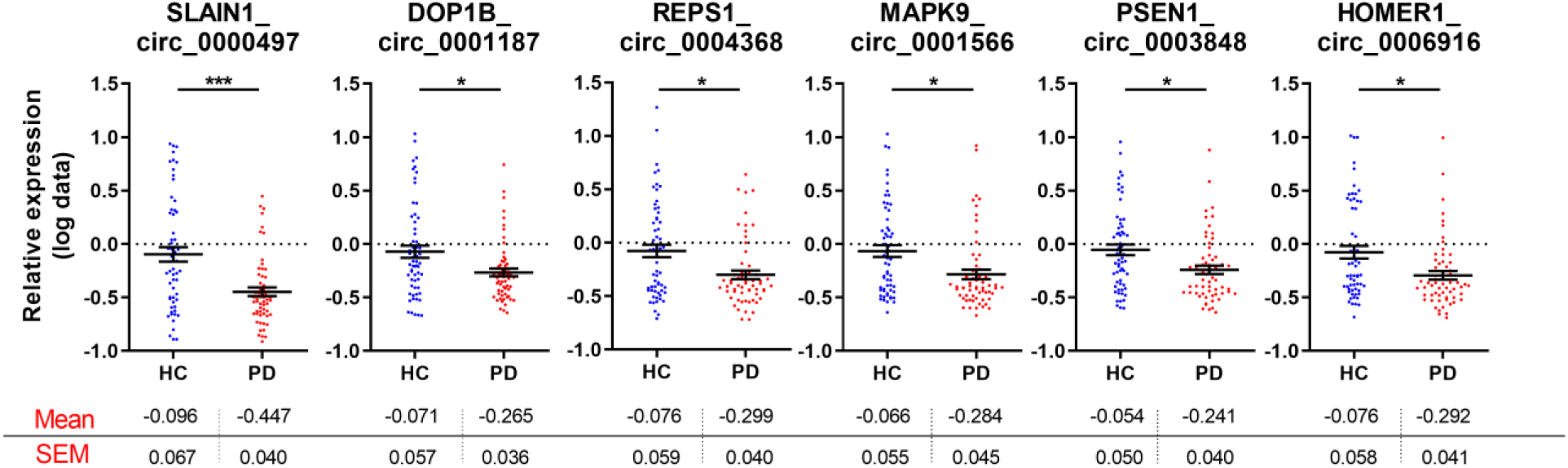
Swarm plots of deregulated circRNAs relative expression in the PBMCs of control and idiopathic PD cohorts. Mean levels +/- SEM are included below each graph. Graphs demonstrate relative expression of log-transformed data. Unpaired t-test was used to determine the significance of differences between the two groups. *p<0.05, **p<0.01, ***p<0.001.

### Association between circRNA levels and clinical features, age or sex

Spearman correlation test was used to relate circRNA levels to iPD patients’ clinical features. We found no correlation between age-at-onset, disease duration, UPDRS III, MMSE, LEDD, H&Y or patients’ on/off state and circRNA levels (Suppl. Table 4 and data not shown). Finally, correcting clinical scores with LEDD did not reveal any more associations (data not shown). In addition, there was no significant correlation between circRNA expression and age or sex in either healthy controls or PD patients.

### Discriminant Analysis

To evaluate the utility of PBMC circRNA levels in discriminating subjects with iPD from healthy controls, ROC curve analysis was performed. The diagnostic sensitivity and specificity of a four circRNA panel (SLAIN1_circ_0000497, SLAIN2_circ_0126525, ANKRD12_circ_0000826, and PSEN1_circ_0003848) were 75.3% (62.1-85.2%) and 78% (65.8-88%), respectively, and the AUC was 0.84 (Fig. 3).

**Figure 3.**
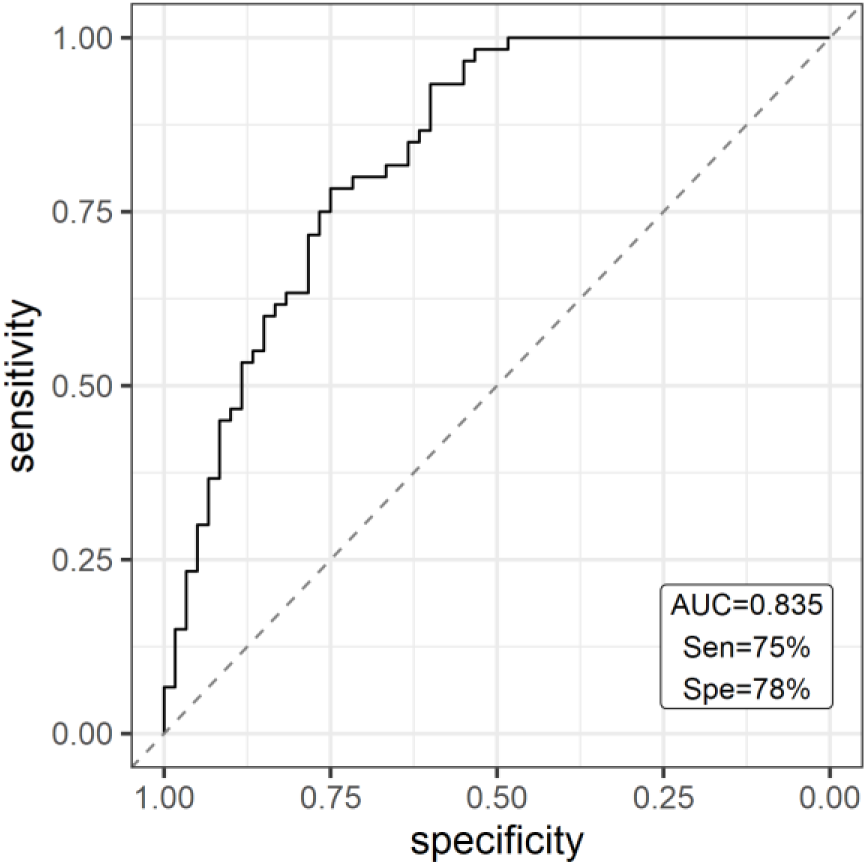
The receiver operating characteristic (ROC) curve analysis for discriminating idiopathic PD from healthy control subjects. ROC curve of four circRNAs (MAPK9_circ_0001566, SLAIN1_circ_0000497, SLAIN2_circ_0126525, and PSEN1_circ_0003848) differentiate iPD from HC cases.

### CeRNA networks

CircRNAs can act as miRNA and RBP sponges for regulating gene expression. To explore the functional role of the deregulated circRNAs we identified all their miRNA and RBP targets. Multiple miRNA binding sites are predicted for each circRNA, with SLAIN1_circ_0000497 and MAPK9_circ_0001566 having the most of MREs (38 and 24, respectively) (Fig. 4A).

**Figure 4.**
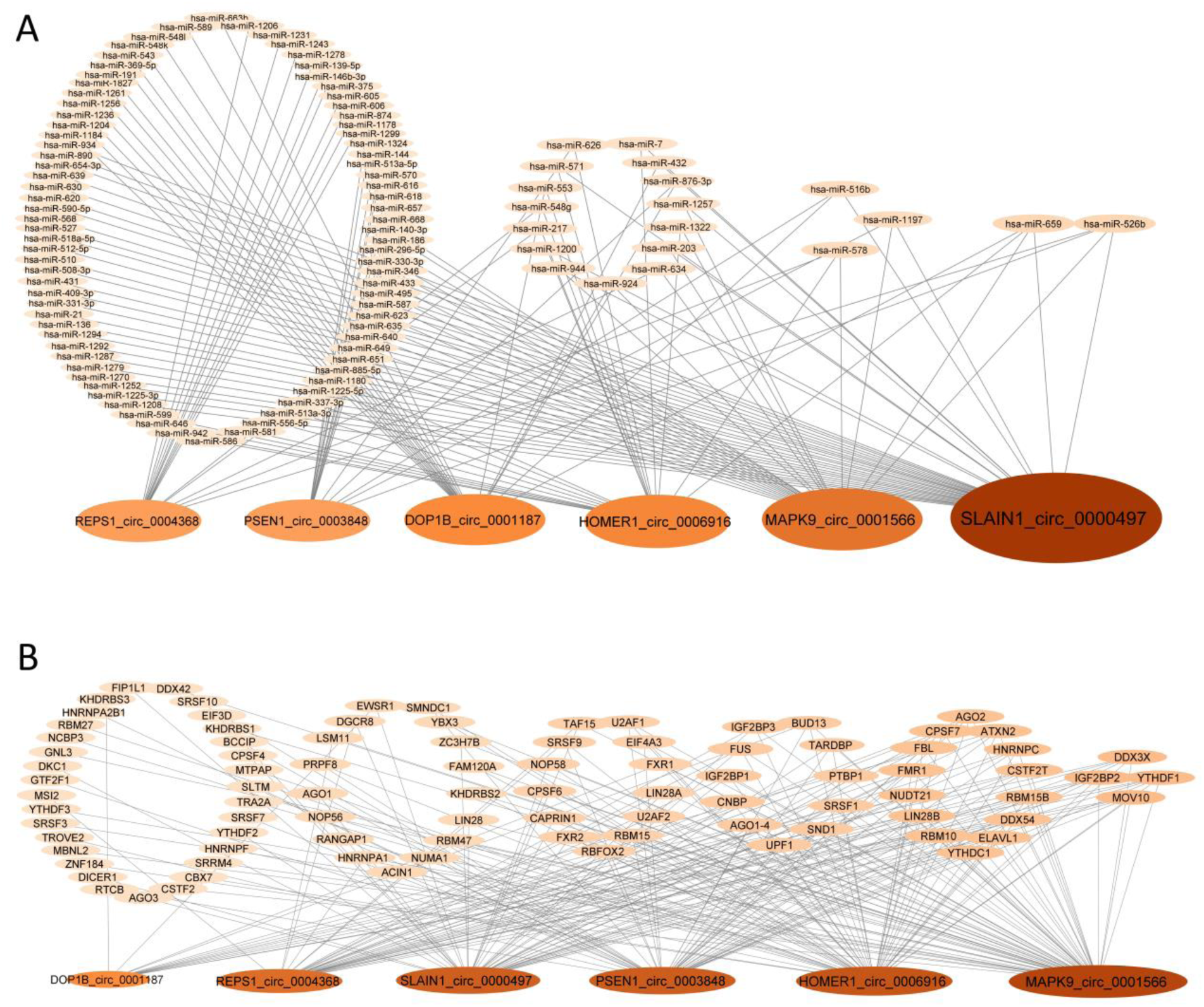
CircRNA target networks. Diagrams show (A) the predicted miRNAs and (B) the CLIP-Seq identified RBPs that bind to differentially expressed circRNAs.

Interestingly, five miRNAs were predicted to be sponged by half or more of the deregulated PD circRNAs. miR-526b and miR-659 are the top targets, sequestered by four deregulated PD circRNAs (Fig. 4A). Fig. 4B shows the deregulated circRNA-RBP network. Like for miRNAs, CLIP-Seq data obtained from StarBase database revealed that the deregulated circRNAs have multiple RBP binding sites. MAPK9_circ_0001566 and HOMER1_circ_0006916 host the most of these sites with 60 and 49 sites, respectively. Interestingly, 29 RBPs were sequestered by four or more deregulated circRNAs.

### CircRNA-miRNA pathway analysis

In order to explore the biological pathways affected by the five miRNAs (miR-516b-5p, miR-526b-5p, miR-578, miR-659-3p, and miR-1197) mostly sequestered by three or more of the deregulated circRNAs, the DIANA mirPath v3 tool was used to align miRNA predicted targets with KEGG pathways and GOslim categories. A Priori gene union analysis of deregulated miRNA targets revealed fourteen KEGG categories as significantly enriched; they included ‘Thyroid hormone signaling pathway’ (p< 0.0015, 24 genes), ‘Regulation of actin cytoskeleton’ (p< 0.015, 42 genes) ‘Phosphatidylinositol signaling pathway’ (p< 0.016, 16 genes), ‘MAPK signaling pathway’ (p< 0.016, 46 genes) and ‘FoxO signaling pathway’ (p< 0.016, 26 genes) (Suppl. Table 5A). Similar findings were obtained using a Posteriori analysis (Suppl. Fig. 2A). Thirty-nine GOslim categories that are controlled by the gene union of the deregulated miRNA targets were enriched following a Priori analysis; these included ‘Cellular protein modification’ (p< 3.93E-19, 283 genes), ‘Nucleic acid binding transcription factor activity’ (p< 7.74E-07, 112 genes), ‘Cytoskeletal protein binding’ (p< 3.91E-09, 102 genes) ‘Cell death’ (p< 7.91E-08, 112 genes), ‘RNA binding’ (p< 6.93E-07, 207 genes) and ‘Response to stress’ (p< 3.72E-05, 225 genes) (Suppl. Table 5B). Similar findings were obtained using a Posteriori analysis (Suppl. Fig. 2B).

## DISCUSSION

We profiled brain-enriched circRNAs in peripheral blood from controls and patients with PD using a RT-qPCR -based approach for three reasons. First, primers could be designed to span the splicing junction which guarantees that only message from the circRNA is amplified. Second, RT-qPCR is the most sensitive method to accurately determine expression changes between cohorts; the alternative microarray approach is prone to errors at multiple levels and nearly always requires a follow up RT-qPCR-based analysis to validate findings. Third, we probed circRNAs that are abundantly expressed in the brain; in this way, we could identify differentially expressed or spliced circRNAs that are more likely associated with the neurological processes of PD.

### Insights into the differentially expressed circRNA genes

We initiated our study with 87 brain-enriched circRNAs from which over half were confidently detected in PBMCs. From these circRNAs, six were differentially expressed in PD with a 17% decrease on average from healthy controls levels. These changes may appear subtle but depending on the circRNA baseline expression levels and considering the relative importance of their multiple targets (transcription factors, RBPs, and miRNAs) as well as the added-up deregulation of the common targets, the biological outcome is expected to be significant.

It has been observed that the biological role of host transcripts reflects on the function of the circRNAs ^17^. We found that the host transcripts of the differentially-expressed circRNAs are not exclusive to brain pathways; rather, they are house-keeping genes, whose functions are best characterized in the CNS as they are essential for neuronal homeostasis. A brief bibliographical overview of their properties is described below.

Hsa_circ_0001566 is hosted by the mitogen-activated protein kinase 9 (MAPK9) gene. MAPK9/JNK2 is a member of the c-Jun n-terminal kinase 1-3 family robustly activated by environmental stresses, including the PD-related neurotoxins lipopolysaccharides (LPS), MPTP and 6-hydroxydopamine (6-OHDA) to mediate neuronal degeneration ^44^. It is indispensable during brain development for neuronal migration, axonal sprouting and guidance, as well as neuronal survival ^45^. Hsa_circ_0006916 is hosted by the homer scaffold protein 1 (HOMER1) gene. HOMER1 is a member of Homer 1-3 family constituting important scaffold proteins at the postsynaptic density that associate with a large number of Ca^2+^-handling proteins, including channels, receptors, and shank scaffolding proteins to regulate intracellular Ca^2+^ homeostasis ^46^. An SNP in the promoter of HOMER1 has been associated with psychotic symptoms in PD ^47^. Further, circHomer1a is reduced in the prefrontal cortex of patients with schizophrenia and bipolar disorder, where it modulates the alternative splicing of mRNA transcripts involved in synaptic plasticity and psychiatric disease ^11^. Hsa_circ_0000497 is hosted by the SLAIN motif family member 1 (SLAIN1) gene. SLAIN1 and 2 are microtubule-associated proteins that promote persistent microtubule growth by recruiting the microtubule polymerase ch-TOG to microtubule plus-ends and thus they are important for axon elongation in developing neurons ^48^. Recently, SLAIN1 was identified as a candidate gene for intellectual disability ^49^. Hsa_circ_0001187 is hosted by the DOP1 leucine zipper like protein B (DOP1B) gene. DOP1B/ DOPEY2/ C21orf5 and its ortholog DOP1A interact with partner MON2 to retrograde transport endosomes from the TGN to the Golgi ^50, 51^. DOP1B is a candidate gene for mental retardation in Down syndrome ^52, 53^ and copy number variations (CNVs) have been observed in AD ^54, 55^. Hsa_circ_0004368 is hosted by the RALBP1 associated eps domain containing 1 (REPS1) gene. REPS1 is a signaling and endocytosis adaptor that interacts with adaptor Intersectin 1 (ITSN1) in clathrin-coated pits and Amphiphysin 1 (AMPH) at the surface of synaptic vesicles ^56^. Mutations in REPS1 are associated with neurodegeneration with brain iron accumulation (NBIA) in the basal ganglia ^57^. Hsa_circ_0003848 is hosted by the presenilin 1 (PSEN1) gene. PSEN1 and its paralog PSEN2 are the endoprotease subunits of the gamma-secretase complex that catalyses the intramembrane cleavage of integral membrane proteins such as Notch receptors and amyloid-beta precursor protein (APP). Mutations in either gene cause early-onset AD ^58^ and α-synuclein accumulation in LB in these patients ^59^. Besides their established role in mediating the formation of Aβ peptide, more recently mutant PS1 has been shown to impair numerous cellular functions such as calcium flux, organization of proteins in different compartments, and protein turnover via vacuolar metabolism ^60^. Interestingly, a novel PSEN1 mutation was recently identified as the likely cause for early-onset Parkinsonism ^61^.

### Correlation between circRNA levels and demographics

There was no significant correlation between the differential expression of a particular circRNA and clinical or demographic measures. Combined interactions with age and sex did not also appear to affect circRNA levels. These findings reinforce current knowledge that the etiology of PD is complex involving a mix of genetic and environmental influences on ageing brain. Similar findings have been observed in miRNA studies ^62^. Further, a pool of four circRNAs discriminated subject with PD from controls with an AUC of 0.84.

### CircRNA-RBP and circRNA-miR interactions

*In silico* approaches were utilized to identify potential biological roles for the deregulated circRNAs by identifying the RBPs and miRNAs that are sequestered preferentially by them. Since the circRNAs were all downregulated, it indicates that target RBP and miRNA functions will be enhanced in PD. The circRNA-RBP network which is based on experimental CLIPS-seq data revealed that 29 RBPs were bound by four or more deregulated circRNAs. Importantly, several of these RBPs are implicated in familial neurodegeneration including Fragile X Mental Retardation Protein 1 (FMR1, Fragile X syndrome and associated disorders), Ataxin 2 (ATXN2, Spinocerebellar ataxia 2, late-onset PD), Fused in Sarcoma (FUS) and TAR DNA binding protein (TARDBP/TDP43) (Amyotrophic lateral sclerosis, Frontotemporal dementia) ^63-72^.

The circRNA-miRNA network revealed five miRNAs that were predicted to be sponged by at least three downregulated PD circRNAs. miR-659-3p is of particular interest as it targets progranulin (PGRN), a neuroprotective and anti-inflammatory protein implicated in FTD ^73-77^. To explore the molecular pathways controlled by the five miRNAs, *in silico* analysis of KEGG pathways and GOslim terms was performed. KEGG categories revealed multiple signaling pathways (Thyroid hormone, Phosphatidylinositol, MAPK, FoxO) implicated in neuronal survival and plasticity and ‘Regulation of actin cytoskeleton’ which is central to pre- and postsynaptic assembly as over-represented ^78-81^. GOslim analysis revealed ‘Cellular protein modification’, ‘Nucleic acid binding transcription factor activity’, ‘Cytoskeletal protein binding’, ‘Cell death’, and ‘Response to stress’ as overrepresented among the biological processes affected. ‘Cellular protein modifications’ such as phosphorylation, ubiquitination, truncation, acetylation, nitration and sumoylation of PD-linked proteins have emerged as important modulators of pathogenic mechanisms in PD ^82, 83^. ‘Transcription factor’ changes indicate that there is not only misexpression at the mRNA translation level by miRNA deregulation, but that there exists a second wave of *en masse* deregulation involving transcription-mediated changes. Finally, deregulation of fine cytoskeletal dynamics is expected to impair trafficking and intracellular signaling pathways and has been recognized as a key insult in the pathogenesis of multiple neurodegenerative diseases including PD ^84, 85^.

## Conclusions

We performed an RT-qPCR-based analysis on RNA extracted from PBMC cells from a cohort of patients with PD and matched controls to identify deregulated circRNAs. The circRNAs investigated are highly-expressed in the human brain. This is the first study of its kind in PD. The measurement of four out of six downregulated circRNAs provided reasonable sensitivity and specificity for PD in this discovery cohort. The circRNAs found deregulated form a robust set of brain-associated circRNAs, that can now be further evaluated, along with other measures, as diagnostic and possible therapeutic targets for PD. *In silico* analysis provided a comprehensive guide of the pathways and processes they control, shedding light on their potential biological role. The impact of these findings will now await further exploration.

## ABBREVIATIONS

6-OHDA: 6-hydroxydopamine
ANCOVA: analysis of covariance
ANOVA: analysis of variance
APP: amyloid-beta precursor protein
AUC: area under the curve
circRNA: circular RNA
CT: computed tomography
DOP1B: DOP1 leucine zipper like protein
FTD: Frontotemporal dementia
GO: gene ontology
H&Y: Hoehn & Yahr
HC: healthy control
HDAC: histone deacetylase
HOMER1: homer scaffold protein 1
hsa: homo sapiens
iPD: idiopathic PD
KEGG: Kyoto encyclopedia of genes and genomes
LEDD: Levodopa equivalent daily dose
LPS: lipopolysaccharides
MAPK9: mitogen-activated protein kinase 9
MMSE: Mini Mental State Examination
MRI: magnetic resonance imaging
NBIA: neurodegeneration with brain iron accumulation
PD: Parkinson’s disease
PSEN1: presenilin 1
REPS1: RALBP1 associated eps domain containing 1
SNCA: alpha-synuclein
SLAIN1: SLAIN motif family member
UPDRS: unified Parkinson’s disease rating scale
TARDBP: TAR DNA Binding Protein
UTR: untranslated region

## ACKNOWLEDGMENTS

The authors are grateful to patients, relatives, and volunteer healthy controls for their participation in this study. This research was financed by Greece and European Union (European Social Fund-ESF) through the Operational Program «Human Resources Development, Education and Lifelong Learning 2014-2020» in the context of the project “Development of diagnostic biomarker tests for Parkinson’s disease” (MIS 5049385). It was also co-financed by the action “Precision medicine Hellenic network in genetic neurodegenerative diseases” (2018ΣE01300001) of the Hellenic public investments program of GSRT and the Michael J. Fox Foundation for Parkinson’s Research (Grant ID 13353).

## AUTHORS’ ROLES

Conceived the study: ED. Neurologically examined patients: AB, NP, LS. Peripheral blood processing: MM. Differential expression analysis: SR, DK, ED. Analyzed data: SR, NP, ED. Bioinformatics analyses: ED. Wrote the manuscript: ED. All authors read, edited and approved the final manuscript.

## SUPPLEMENTAL FIGURES

**Supplemental Figure 1.**
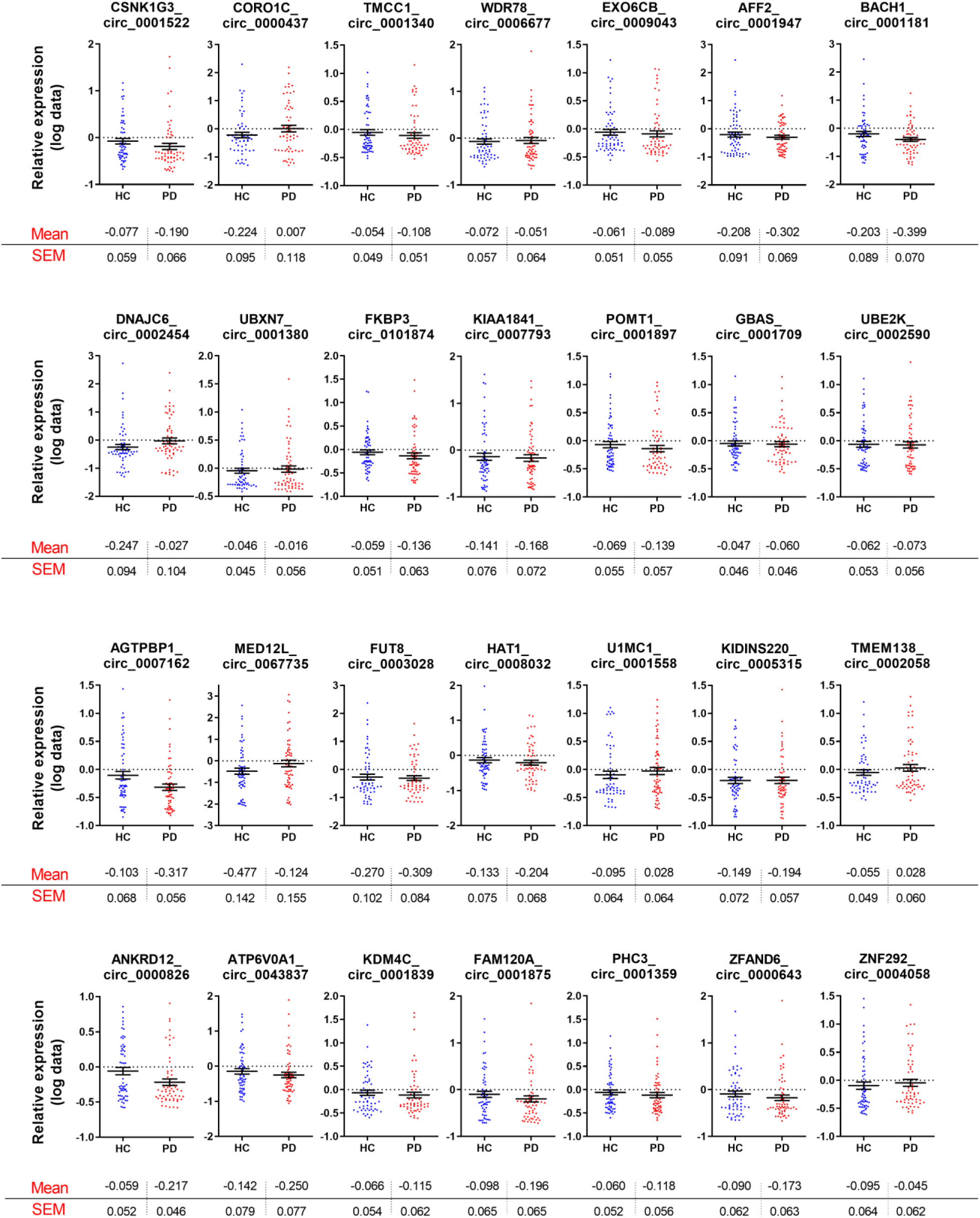
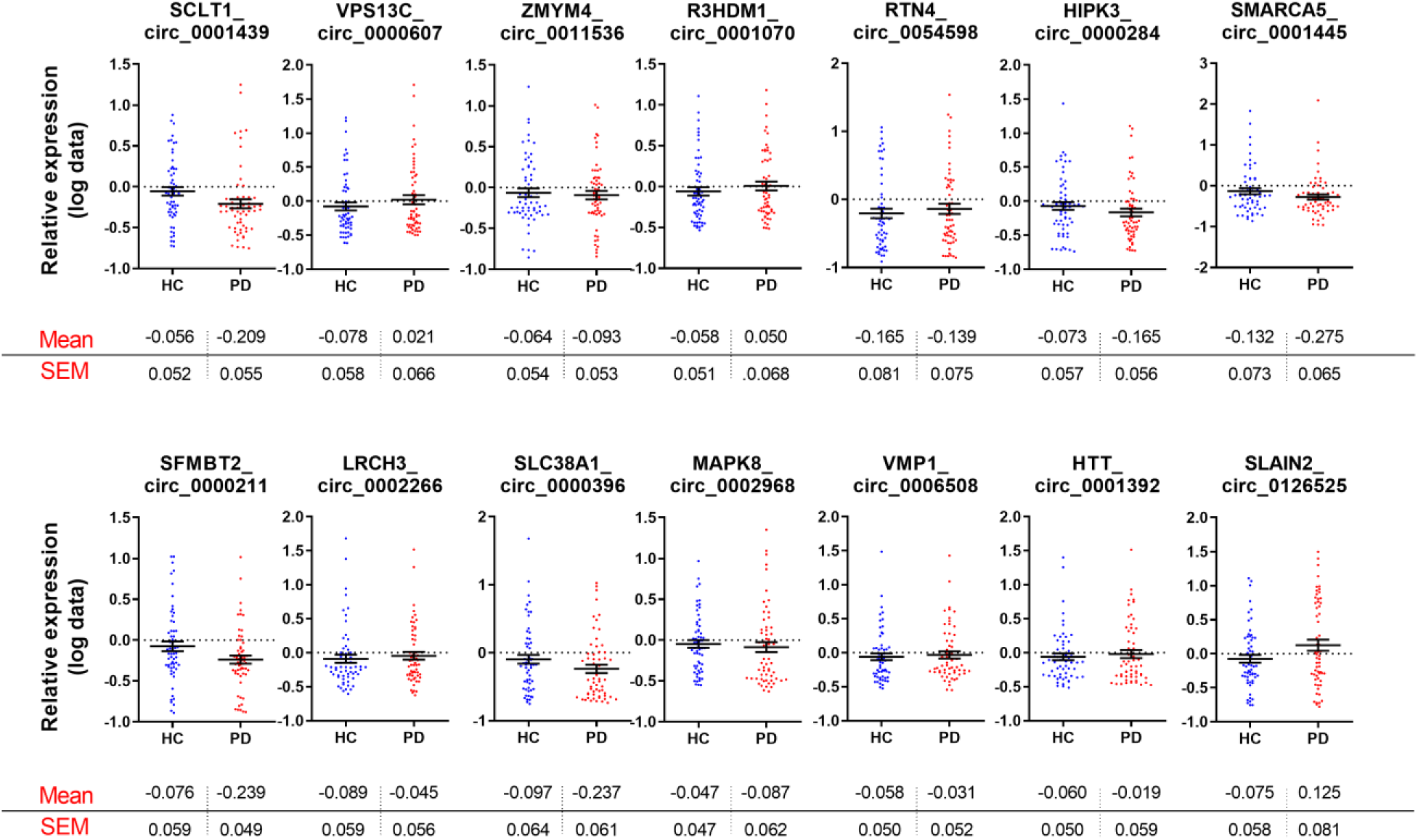
Swarm plots for the 42 circRNAs whose relative expression is not significantly altered in the PBMCs of idiopathic PD patients. Mean levels +/- SEM are included below each graph. Graphs demonstrate relative expression of log-transformed data. Unpaired t-test was used to determine the significance of differences between the two groups. *p<0.05, **p<0.01, ***p<0.001.

**Supplemental Figure 2.**
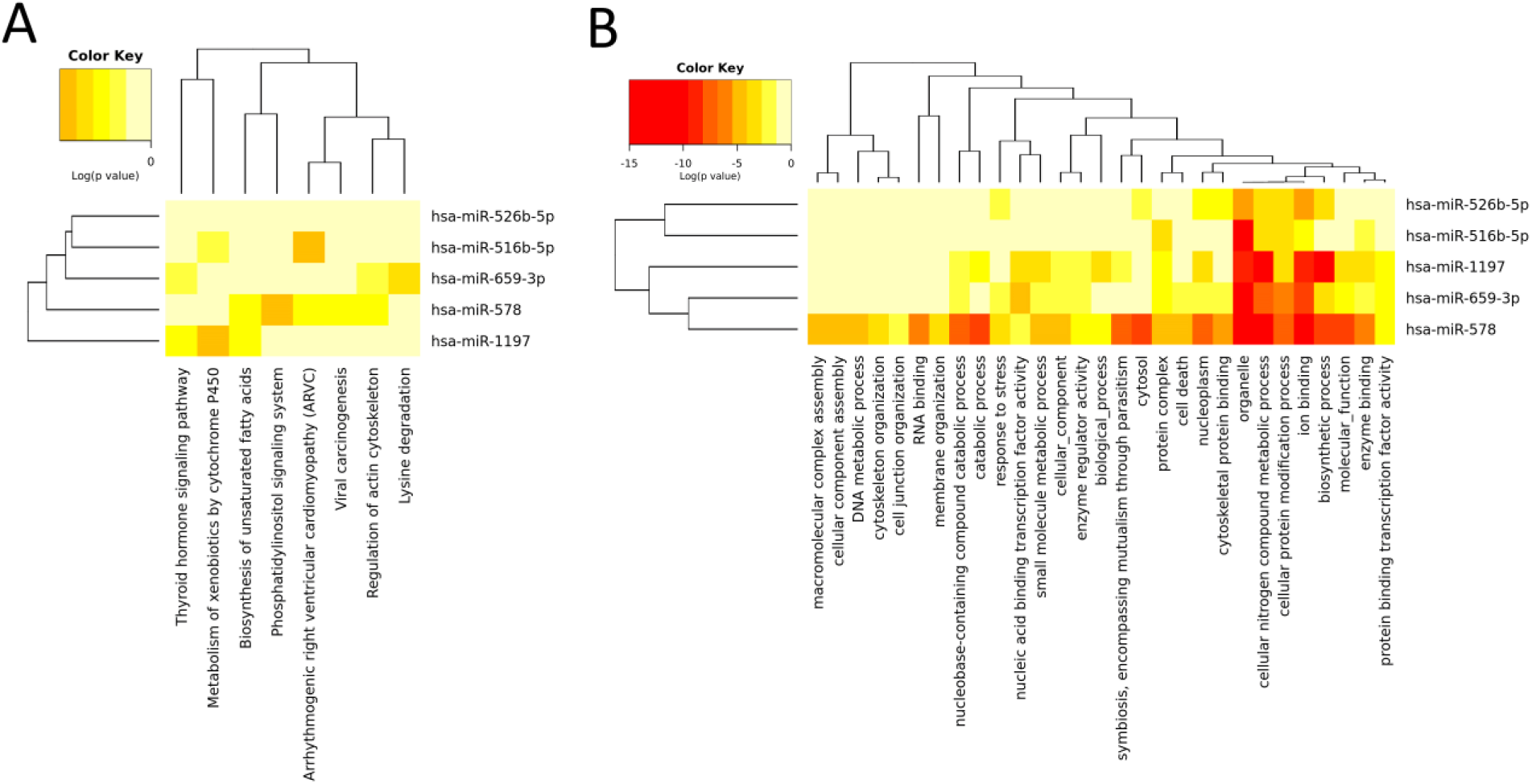
KEGG and GOslim classifications of the circRNA-miRNA target genes. **(A) KEGG and (B) GOslim categories** union of the RNA targets of the five miRNAs sequestered by three or more of the deregulated PD circRNAs (miR-516b-5p, miR-526b-5p, miR-578, miR-659-3p, miR-1197). They were prepared using the DIANA-miRPath v3.0 interface using default values (p-value threshold 0.05, microT-CDS threshold 0.8).

## SUPPLEMENTAL TABLES

**Supplemental Table 1.**
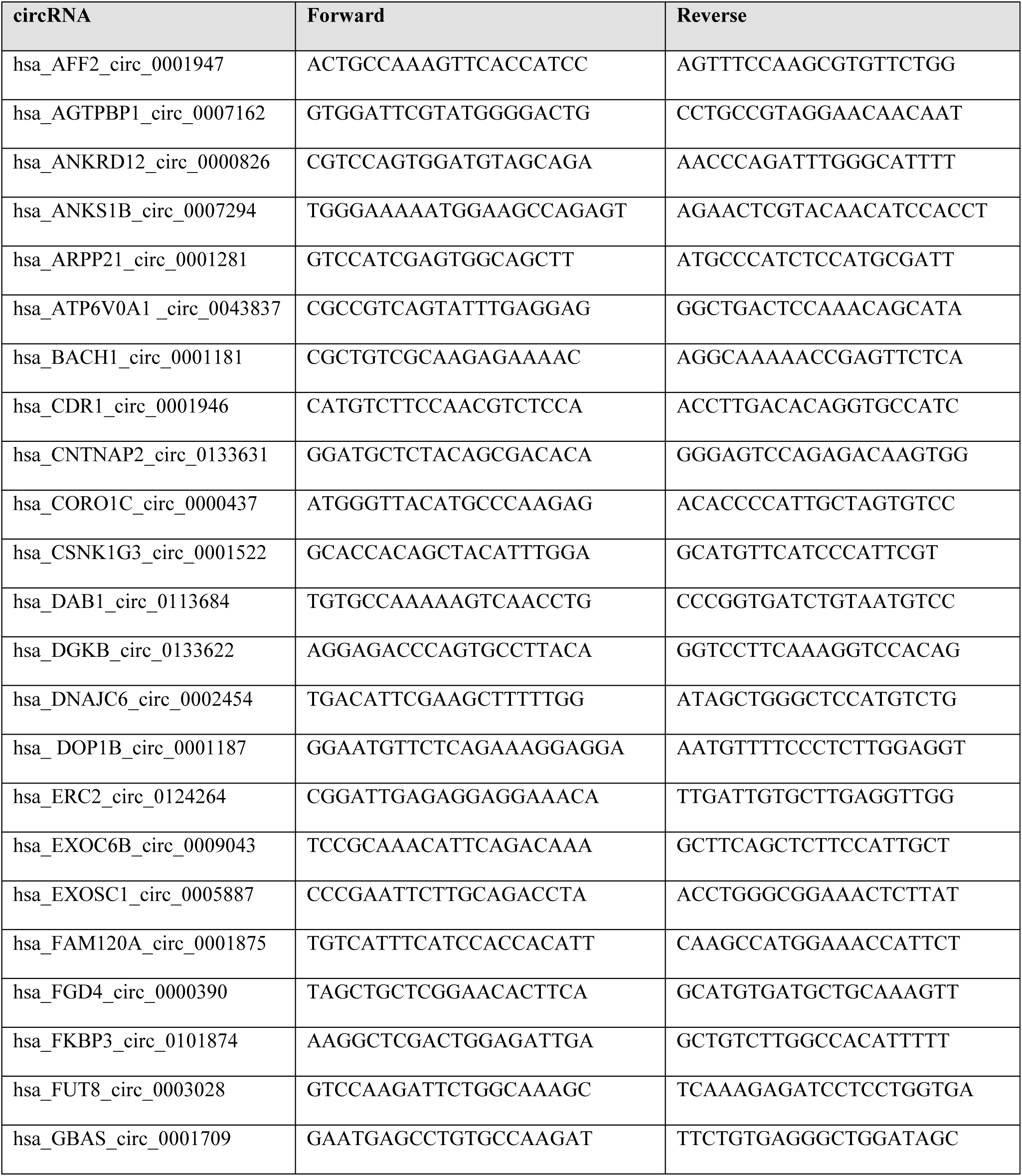
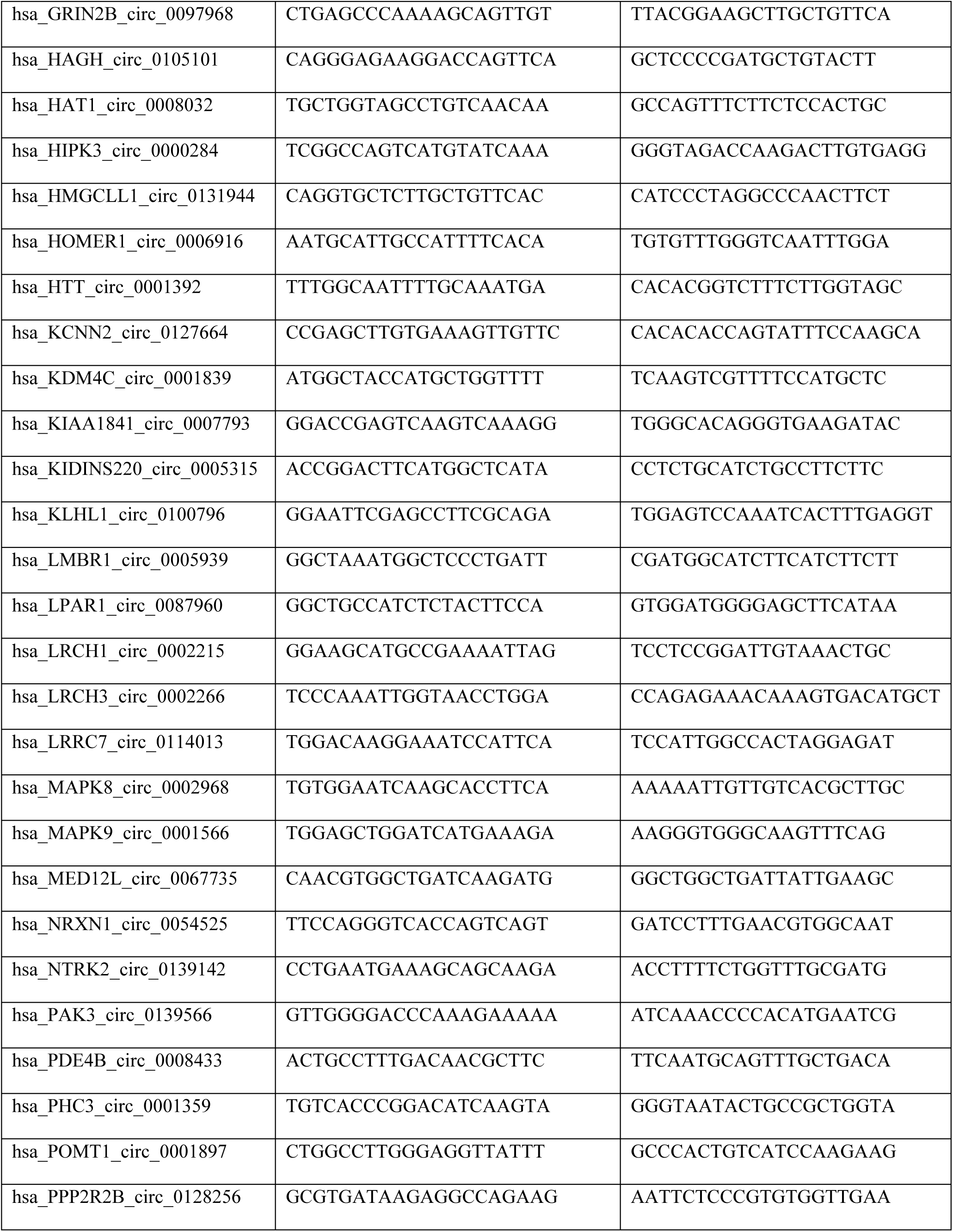
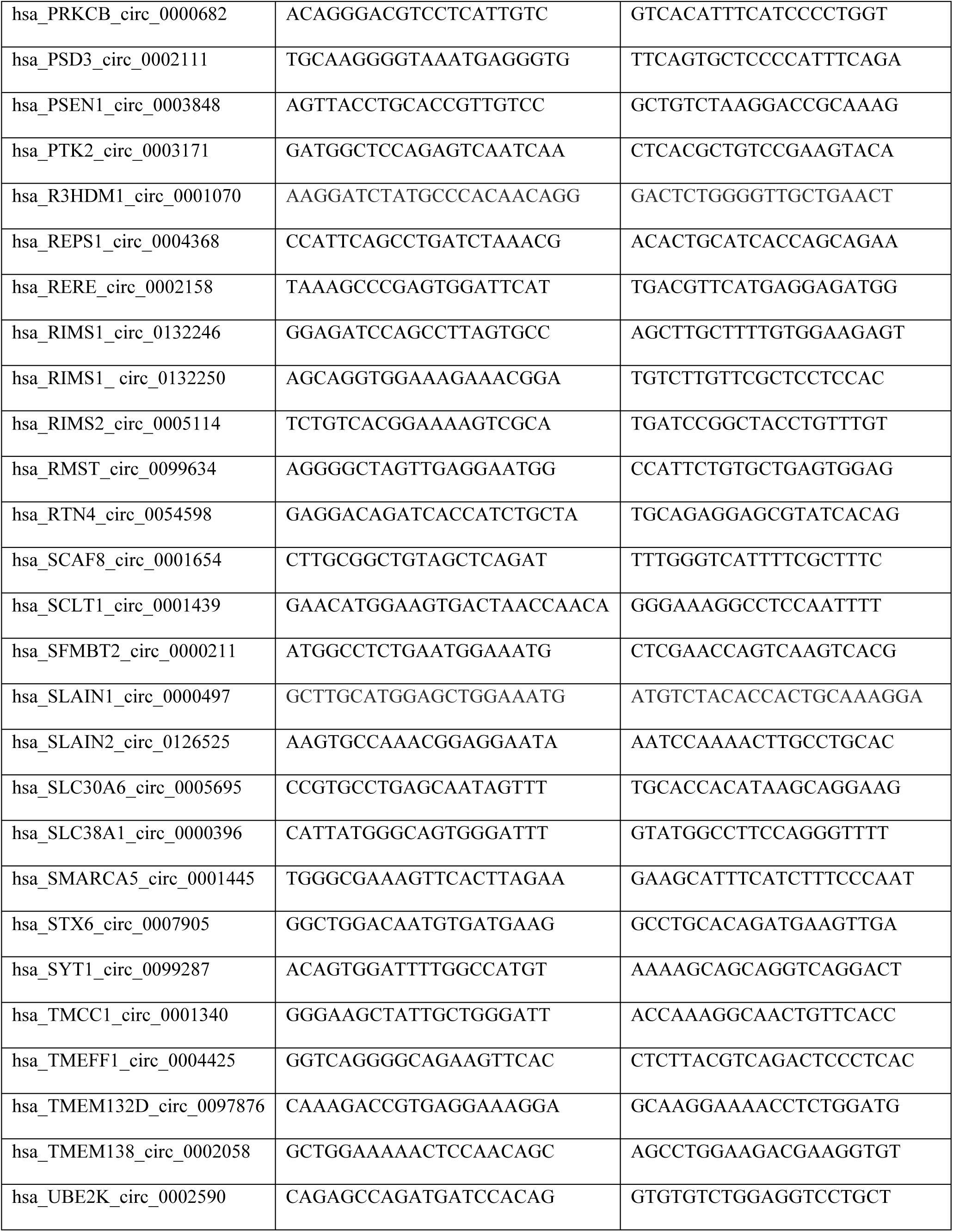
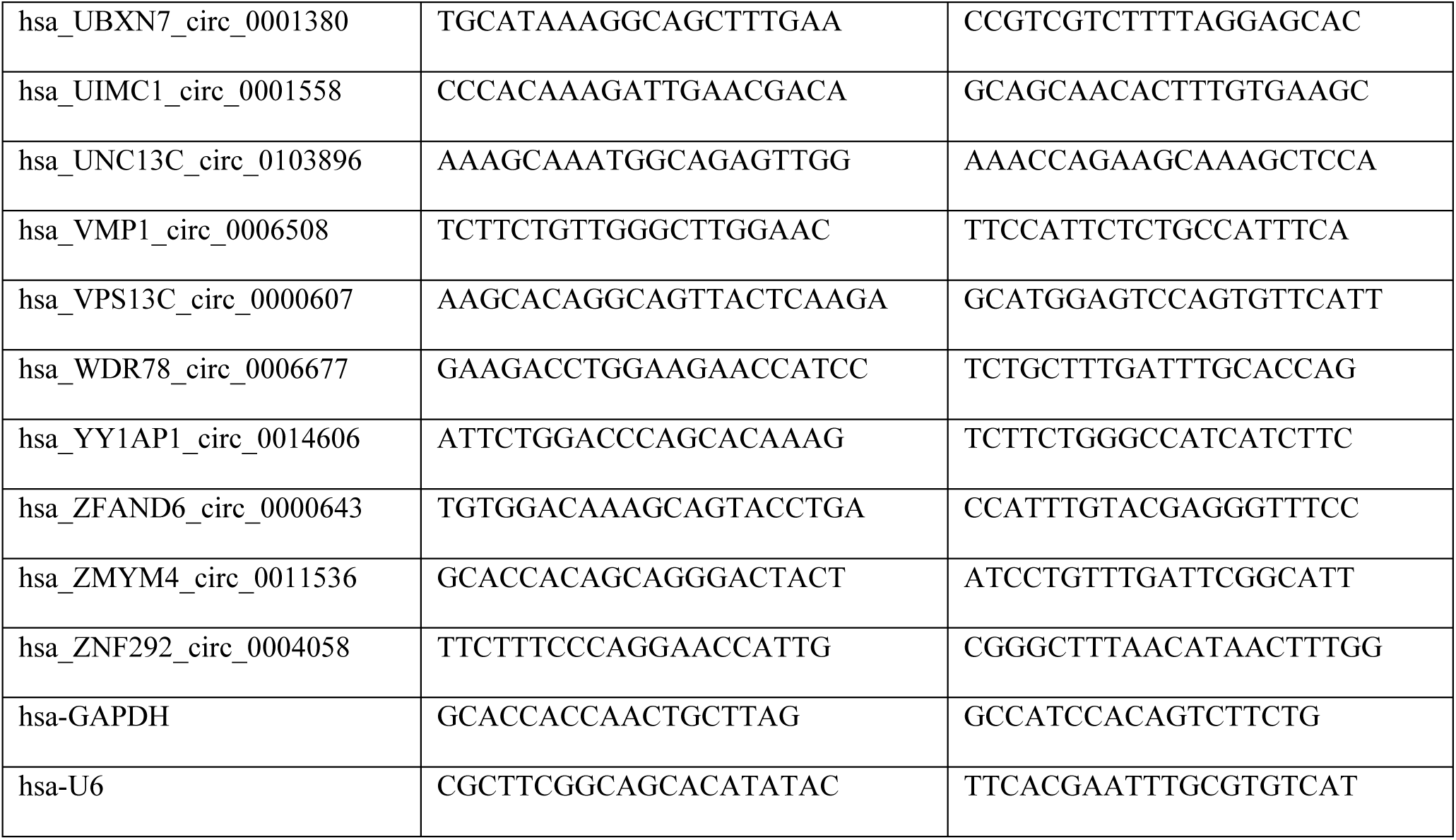
Primer sequences used for real-time PCR.

**Supplemental Table 2.** List of 39 circRNAs not detected in PBMCs.

|  |  |  |
| --- | --- | --- |
| ANKS1B_circ_0007294 | KLHL1_circ_0100796 | RERE_circ_0002158 |
| ARPP21_circ_0001281 | LMBR1_circ_0005939 | RIMS1_circ_0132246 |
| CDR1_circ_0001946 | LPAR1_circ_0087960 | RIMS1_circ_0132250 |
| CNTNAP2_circ_0133631 | LRCH1_circ_0002215 | <u>RIMS2_circ_0005114</u> |
| DAB1_circ_0113684 | LRRC7_circ_0114013 | RMST_circ_0099634 |
| DGKB_circ_0133622 | NRXN1_circ_0054525 | SCAF8_circ_0001654 |
| ERC2_circ_0124264 | NTRK2_circ_0139142 | SLC30A6_circ_0005695 |
| EXOSC1_circ_0005887 | PAK3_circ_0139566 | STX6_circ_0007905 |
| FGD4_circ_0000390 | PDE4B_circ_0008433 | SYT1_circ_0099287 |
| GRIN2B_circ_0097968 | PPP2R2B_circ_0128256 | TMEFF1_circ_0004425 |
| HAGH_circ_0105101 | PRKCB_circ_0000682 | TMEM132D_circ_0097876 |
| HMGCLL1_circ_0131944 | PSD3_circ_0002111 | UNC13C_circ_0103896 |
| KCNN2_circ_0127664 | PTK2_circ_0003171 | YY1AP1_circ_0014606 |

**Supplemental Table 3.** Basic characteristics and comparative PBMC circRNA expression in idiopathic PD patients and healthy controls. CircRNA size and expression in the brain as well as host transcript expression in the body. Means with their respective standard deviation (std) for both groups are shown. Statistically significant differences in comparison to healthy controls (unpaired t-test) are highlighted in grey. Multiple comparison analysis (adjusted *P* values) calculated according to Benjamini-Hochberg false discovery rate method.

| circRNA ID | Spliced Seq length | Expression score in the human brain* | Host gene symbol | Host gene mRNA expression in the human body** | Mean (± std) | | Unpaired t-test *P* value | Adjusted *P* values |
| --- | --- | --- | --- | --- | --- | --- | --- | --- |
|  |  |  |  |  | HC | iPD |  |  |
| circ_0001380 | 247 | 15326 | UBXN7 | Ubiquitous, expressed least in the brain except cerebellum where expression is similar to other tissues | -0.046<br>(0.348) | -0.016<br>(0.432) | 0.678 | 0.812 |
| circ_0000284 | 1099 | 13889 | HIPK3 | Ubiquitous, expressed least in the brain | -0.073<br>(0.443) | -0.165<br>(0.435) | 0.252 | 0.603 |
| circ_0001445 | 269 | 9354 | SMARCA5 | Ubiquitous, lower expression in the brain | -0.132<br>(0.565) | -0.275<br>(0.504) | 0.147 | 0.415 |
| circ_0009043 | 390 | 9121 | EXOC6B | Ubiquitous, enriched in the skin | -0.061<br>(0.395) | -0.089<br>(0.425) | 0.701 | 0.812 |
| circ_0001947 | 861 | 9058 | AFF2 | Ubiquitous, highly enriched in the cerebellum | -0.208<br>(0.702) | -0.302<br>(0.531) | 0.412 | 0.706 |
| circ_0002590 | 336 | 8841 | UBE2K | Ubiquitous, enriched in the cerebellum | -0.062<br>(0.409) | -0.073<br>(0.437) | 0.895 | 0.895 |
| circ_0000437 | 251 | 8402 | CORO1C | Ubiquitous, expressed least in the brain | -0.224<br>(0.736) | 0.007<br>(0.911) | 0.130 | 0.390 |
| circ_0000497 | 788 | 6004 | SLAIN1 | Brain-specific | -0.096<br>(0.520) | -0.447<br>(0.313) | <0.0001<br>*** | 0.001*** |
| circ_0054598 | 2457 | 5985 | RTN4 | Ubiquitous, enriched in the brain | -0.165<br>(0.624) | -0.139<br>(0.583) | 0.815 | 0.850 |
| circ_0001522 | 536 | 5649 | CSNK1G3 | Ubiquitous, lower expression in the brain | -0.077<br>(0.452) | -0.190<br>(0.505) | 0.205 | 0.518 |
| circ_0001070 | 307 | 5300 | R3HDM1 | Brain-specific | -0.058<br>(0.394) | 0.050<br>(0.523) | 0.204 | 0.518 |
| circ_0011536 | 755 | 4662 | ZMYM4 | Ubiquitous, expressed least in the brain except cerebellum where expression is somewhat higher than in other tissues | -0.064<br>(0.417) | -0.093<br>(0.406) | 0.694 | 0.812 |
| circ_0000607 | 606 | 4056 | VPS13C | Ubiquitous, expressed least in the brain except cerebellum where expression is similar to other tissues | -0.078<br>(0.449) | 0.021<br>(0.514) | 0.266 | 0.603 |
| circ_0001439 | 396 | 3870 | SCLT1 | Ubiquitous, expressed least in the brain except cerebellum where expression is somewhat higher than in other tissues | -0.056<br>(0.399) | -0.209<br>(0.429) | 0.046* | 0.210 |
| circ_0000826 | 994 | 3868 | ANKRD12 | Ubiquitous, expressed least in the brain except cerebellum where expression is somewhat higher than in other tissues | -0.059<br>(0.405) | -0.217<br>(0.355) | 0.024* | 0.142 |
| circ_0000211 | 434 | 3783 | SFMBT2 | Ubiquitous | -0.076<br>(0.457) | -0.239<br>(0.381) | 0.036* | 0.189 |
| circ_0002266 | 378 | 3204 | LRCH3 | Ubiquitous, expressed least in the brain except cerebellum where expression is somewhat higher than in other tissues | -0.089<br>(0.458) | -0.045<br>(0.433) | 0.595 | 0.782 |
| circ_0002968 | 499 | 3123 | MAPK8 | Ubiquitous, highly enriched in the cerebellum | -0.047<br>(0.361) | -0.087<br>(0.476) | 0.603 | 0.782 |
| circ_0000396 | 522 | 3086 | SLC38A1 | Ubiquitous | -0.097<br>(0.498) | -0.237<br>(0.472) | 0.115 | 0.380 |
| circ_0001181 | 1836 | 3082 | BACH1 | ubiquitous, expressed least in the brain | -0.203<br>(0.691) | -0.399<br>(0.538) | 0.086 | 0.344 |
| circ_0006916 | 522 | 3026 | HOMER1 | Ubiquitous, enriched in the brain | -0.076<br>(0.452) | -0.292<br>(0.316) | 0.003** | 0.036* |
| circ_0001709 | 212 | 2889 | GBAS | Ubiquitous, highly enriched in the skeletal muscle | -0.047<br>(0.354) | -0.060<br>(0.359) | 0.839 | 0.857 |
| circ_0001897 | 158 | 2882 | POMT1 | Ubiquitous, highly enriched in the cerebellum and testis | -0.069<br>(0.426) | -0.139<br>(0.439) | 0.375 | 0.667 |
| circ_0101874 | 502 | 2687 | FKBP3 | Ubiquitous, enriched in the brain | -0.059<br>(0.395) | -0.136<br>(0.482) | 0.347 | 0.640 |
| circ_0002454 | 350 | 2511 | DNAJC6 | Brain-specific | -0.247<br>(0.728) | -0.027<br>(0.806) | 0.119 | 0.380 |
| circ_0001558 | 397 | 2476 | UIMC1 | Ubiquitous, expressed least in the brain except cerebellum where expression is similar to other tissues | -0.095<br>(0.498) | 0.028<br>(0.497) | 0.459 | 0.711 |
| circ_0001187 | 301 | 2263 | DOP1B | Ubiquitous, highly enriched in the cerebellum | -0.071<br>(0.440) | -0.265<br>(0.282) | 0.005** | 0.038* |
| circ_0006508 | 440 | 2229 | VMP1 | Ubiquitous, expressed least in the brain | -0.058<br>(0.384) | -0.031<br>(0.402) | 0.711 | 0.812 |
| circ_0006677 | 473 | 2190 | WDR78 | Testis, lung | -0.072<br>(0.439) | -0.051<br>(0.492) | 0.801 | 0.850 |
| circ_0008032 | 302 | 2153 | HAT1 | Ubiquitous, expressed least in the brain | -0.133<br>(0.579) | -0.204<br>(0.526) | 0.482 | 0.723 |
| circ_0043837 | 444 | 1892 | ATP6V0A1 | Ubiquitous, highly enriched in the brain | -0.142<br>(0.611) | -0.250<br>(0.597) | 0.334 | 0.640 |
| circ_0004368 | 352 | 1882 | REPS1 | Ubiquitous, highly enriched in the cerebellum | -0.076<br>(0.455) | -0.299<br>(0.313) | 0.002** | 0.036* |
| circ_0005315 | 308 | 1820 | KIDINS220 | Ubiquitous, enriched in the cerebellum | -0.149<br>(0.554) | -0.194<br>(0.438) | 0.628 | 0.793 |
| circ_0002058 | 248 | 1775 | TMEM138 | ubiquitous, expressed least in the brain | -0.055<br>(0.379) | 0.028<br>(0.465) | 0.286 | 0.603 |
| circ_0001566 | 497 | 1745 | MAPK9 | Ubiquitous, enriched in the brain | -0.066<br>(0.425) | -0.284<br>(0.352) | 0.002** | 0.036* |
| circ_0007793 | 284 | 1645 | KIAA1841 | Ubiquitous, somewhat brain-enriched | -0.141<br>(0.591) | -0.168<br>(0.556) | 0.793 | 0.850 |
| circ_0001340 | 251 | 1631 | TMCC1 | Ubiquitous, enriched in the cerebellum | -0.054<br>(0.380) | -0.108<br>(0.389) | 0.443 | 0.711 |
| circ_0007162 | 1121 | 1526 | AGTPBP1 | Ubiquitous, somewhat brain-enriched | -0.103<br>(0.524) | -0.317<br>(0.434) | 0.017* | 0.113 |
| circ_0003848 | 222 | 1430 | PSEN1 | Ubiquitous, highly enriched in the spinal cord | -0.054<br>(0.388) | -0.241<br>(0.308) | 0.004** | 0.038* |
| circ_0004058 | 370 | 1422 | ZNF292 | Ubiquitous, expressed least in the brain except cerebellum where expression is similar to other tissues | -0.095<br>(0.491) | -0.045<br>(0.479) | 0.577 | 0.782 |
| circ_0001875 | 556 | 1345 | FAM120A | Ubiquitous, expressed least in the brain | -0.098<br>(0.504) | -0.196<br>(0.500) | 0.289 | 0.603 |
| circ_0003028 | 430 | 1336 | FUT8 | Ubiquitous, highly enriched in the spinal cord | -0.270<br>(0.792) | -0.309<br>(0.654) | 0.850 | 0.850 |
| circ_0001839 | 292 | 1280 | KDM4C | Ubiquitous, highly enriched in the cerebellum | -0.066<br>(0.418) | -0.115<br>(0.482) | 0.557 | 0.782 |
| circ_0001392 | 484 | 1270 | HTT | Ubiquitous, enriched in the cerebellum | -0.060<br>(0.384) | -0.019<br>(0.453) | 0.602 | 0.782 |
| circ_0067735 | 457 | 952 | MED12L | Adrenal glands, brain, cervix, pituitary and testis | -0.477<br>(1.100) | -0.124<br>(1.202) | 0.096 | 0.354 |
| circ_0000643 | 381 | 761 | ZFAND6 | Ubiquitous, expressed least in the brain | -0.090<br>(0.477) | -0.173<br>(0.467) | 0.341 | 0.640 |
| circ_0126525 | 971 | 720 | SLAIN2 | Ubiquitous, expressed least in the brain | -0.075<br>(0.450) | 0.125<br>(0.628) | 0.048* | 0.210 |
| circ_0001359 | 505 | 709 | PHC3 | Ubiquitous, expressed least in the brain | -0.060<br>(0.403) | -0.118<br>(0.434) | 0.454 | 0.711 |

\*According to Rybak et al.
\*\* According to GTEx portal.

**Supplemental Table 4.** Correlation between relative circRNA expression and sex, age, age-at-onset, PD duration, HY, UPDRS and MMSE scores and LEDD of PD patients.

| circRNA | Sex<br>U score<br>(*P* value) | Age<br>Rho<br>(*P* value) | Age-at-onset<br>Rho<br>(*P* value) | PD<br>duration<br>Rho<br>(*P* value) | UPDRS<br>III<br>Rho<br>(*P* value) | MMSE<br>Rho<br>(*P* value) | HY<br>Rho<br>(*P* value) | LEDD<br>Rho<br>(*P* value) |
| --- | --- | --- | --- | --- | --- | --- | --- | --- |
| UBXN7_circ<br>_0001380 | 0.027<br>(0.8684) | -0.012<br>(0.8949) | -0.142<br>(0.2831) | 0.041<br>(0.7579) | 0.143<br>(0.2788) | 0.096<br>(0.4674) | 0.113<br>(0.3953) | 0.055<br>(0.6791) |
| HIPK3_circ<br>_0000284 | 1.694<br>(0.1930) | 0.004<br>(0.9627) | -0.129<br>(0.3266) | -0.063<br>(0.6303) | -0.036<br>(0.7847) | 0.090<br>(0.4924) | -0.052<br>(0.6943) | 0.100<br>(0.4450) |
| SMARCA5_circ<br>_0001445 | 0.778<br>(0.3776) | -0.035<br>(0.7078) | -0.119<br>(0.3663) | -0.174<br>(0.1847) | 0.018<br>(0.8931) | 0.082<br>(0.5321) | 0.005<br>(0.9704) | 0.006<br>(0.9643) |
| EXO6CB_circ<br>_0009043 | 1.129<br>(0.2880) | 0.142<br>(0.1215) | 0.225<br>(0.0836) | -0.116<br>(0.3793) | -0.014<br>(0.9165) | -0.021<br>(0.8736) | -0.066<br>(0.6187) | -0.024<br>(0.8551) |
| AFF2_circ<br>_0001947 | 0.011<br>(0.9159) | 0.107<br>(0.2446) | 0.101<br>(0.4418) | -0.036<br>(0.7871) | 0.050<br>(0.7055) | -0.048<br>(0.7130) | -0.138<br>(0.2917) | 0.156<br>(0.2334) |
| UBE2K_circ<br>_0002590 | 0.010<br>(0.9208) | -0.035<br>(0.7022) | -0.143<br>(0.2773) | -0.187<br>(0.1535) | -0.167<br>(0.2024) | 0.065<br>(0.6205) | -0.012<br>(0.9295) | -0.139<br>(0.2898) |
| CORO1C_circ<br>_0000437 | 0.035<br>(0.8521) | -0.089<br>(0.3313) | -0.390<br>(0.0021) | 0.211<br>(0.1050) | 0.090<br>(0.4928) | 0.039<br>(0.7691) | 0.019<br>(0.8856) | 0.183<br>(0.1618) |
| SLAIN1_circ<br>_0000497 | 0.026<br>(0.8717) | -0.098<br>(0.2870) | -0.085<br>(0.5188) | -0.109<br>(0.4071) | 0.094<br>(0.4771) | 0.257<br>(0.0472) | 0.132<br>(0.3146) | 0.050<br>(0.7061) |
| RTN4_circ<br>_0054598 | 3.005<br>(0.0830) | 0.035<br>(0.7015) | -0.203<br>(0.1191) | -0.073<br>(0.5807) | -0.033<br>(0.8049) | 0.077<br>(0.5578) | -0.016<br>(0.9048) | 0.027<br>(0.8353) |
| CSNK1G3_circ<br>_0001522 | 3.347<br>(0.0673) | -0.049<br>(0.5978) | 0.083<br>(0.5300) | -0.217<br>(0.0987) | -0.018<br>(0.8931) | 0.172<br>(0.1924) | -0.088<br>(0.5066) | -0.115<br>(0.3837) |
| R3HDM1_circ<br>_0001070 | 0.078<br>(0.7798) | 0.010<br>(0.9160) | 0.033<br>(0.8001) | -0.127<br>(0.3322) | -0.224<br>(0.0850) | 0.061<br>(0.6445) | -0.156<br>(0.2326) | -0.153<br>(0.2434) |
| ZMYM4_circ<br>_0011536 | 3.556<br>(0.0593) | -0.085<br>(0.3584) | -0.044<br>(0.7413) | -0.341<br>(0.0078) | -0.123<br>(0.3479) | 0.122<br>(0.3540) | 0.000<br>(0.9976) | -0.081<br>(0.5403) |
| VPS13C_circ<br>_0000607 | 0.001<br>(0.9802) | 0.049<br>(0.5951) | 0.018<br>(0.8923) | -0.048<br>(0.7134) | -0.073<br>(0.5790) | -0.204<br>(0.1181) | 0.090<br>(0.4930) | -0.058<br>(0.6610) |
| SCLT1_circ<br>_0001439 | 0.028<br>(0.8668) | 0.016<br>(0.8636) | -0.159<br>(0.2264) | -0.037<br>(0.7774) | 0.094<br>(0.4769) | 0.048<br>(0.7169) | 0.055<br>(0.6761) | 0.184<br>(0.1589) |
| ANKRD12_<br>circ_0000826 | 0.285<br>(0.5931) | 0.008<br>(0.9346) | -0.091<br>(0.4876) | 0.016<br>(0.9014) | 0.090<br>(0.4944) | -0.023<br>(0.8612) | 0.165<br>(0.2071) | 0.116<br>(0.3755) |
| SFMBT2_circ<br>_0000211 | 0.519<br>(0.4711) | 0.100<br>(0.2778) | 0.008<br>(0.9543) | -0.114<br>(0.3873) | 0.053<br>(0.6898) | 0.244<br>(0.0598) | 0.092<br>(0.4864) | 0.003<br>(0.9828) |
| LRCH3_circ<br>_0002266 | 0.556<br>(0.4559) | -0.042<br>(0.6471) | -0.094<br>(0.4739) | -0.236<br>(0.0692) | -0.104<br>(0.4283) | 0.087<br>(0.5074) | 0.070<br>(0.5965) | -0.061<br>(0.6453) |
| MAPK8_circ<br>_0002968 | 0.846<br>(0.3578) | -0.064<br>(0.4898) | 0.037<br>(0.7785) | -0.163<br>(0.2146) | 0.040<br>(0.7634) | 0.096<br>(0.4648) | 0.115<br>(0.3814) | -0.090<br>(0.4943) |
| SLC38A1_<br>circ_0000396 | 0.062<br>(0.8037) | -0.111<br>(0.2284) | 0.207<br>(0.1124) | -0.149<br>(0.2565) | 0.024<br>(0.8558) | 0.087<br>(0.5068) | 0.004<br>(0.9766) | -0.135<br>(0.3040) |
| BACH1_circ<br>_0001181 | 3.978<br>(0.0461) | -0.133<br>(0.1488) | -0.110<br>(0.4034) | -0.028<br>(0.8333) | 0.070<br>(0.5948) | 0.070<br>(0.5932) | 0.066<br>(0.6146) | 0.254<br>(0.0498) |
| HOMER1_<br>circ_0006916 | 0.102<br>(0.7490) | -0.112<br>(0.2253) | -0.040<br>(0.7623) | -0.090<br>(0.4928) | 0.099<br>(0.4499) | 0.068<br>(0.6068) | 0.157<br>(0.2318) | 0.032<br>(0.8105) |
| GBAS_circ<br>_0001709 | 0.892<br>(0.3450) | -0.041<br>(0.6604) | -0.066<br>(0.6138) | -0.001<br>(0.9951) | 0.111<br>(0.4003) | 0.201<br>(0.1244) | 0.185<br>(0.1566) | 0.025<br>(0.8487) |
| POMT1_circ<br>_0001897 | 1.293<br>(0.2555) | 0.032<br>(0.7285) | 0.002<br>(0.9851) | -0.130<br>(0.3239) | -0.006<br>(0.9630) | -0.095<br>(0.4723) | 0.012<br>(0.9301) | -0.011<br>(0.9336) |
| FKBP3_circ<br>_0101874 | 0.030<br>(0.8633) | -0.073<br>(0.4314) | -0.009<br>(0.9472) | -0.142<br>(0.2839) | 0.104<br>(0.4318) | 0.111<br>(0.4040) | 0.016<br>(0.9041) | 0.050<br>(0.7091) |
| DNAJC6_circ<br>_0002454 | 0.575<br>(0.4485) | -0.107<br>(0.2443) | -0.379<br>(0.0028) | 0.000<br>(0.9985) | -0.011<br>(0.9352) | 0.019<br>(0.8864) | -0.008<br>(0.9514) | 0.078<br>(0.5516) |
| U1MC1_circ<br>_0001558 | 2.384<br>(0.1226) | -0.010<br>(0.9113) | -0.070<br>(0.5960) | -0.223<br>(0.0868) | -0.181<br>(0.1657) | 0.083<br>(0.5273) | -0.104<br>(0.4310) | -0.035<br>(0.7909) |
| DOP1B_circ<br>_0001187 | 0.812<br>(0.3676) | -0.035<br>(0.7063) | 0.020<br>(0.8778) | -0.101<br>(0.4416) | -0.147<br>(0.2623) | -0.075<br>(0.5711) | -0.156<br>(0.2354) | -0.267<br>(0.0396) |
| VMP1_circ<br>_0006508 | 0.785<br>(0.3756) | -0.071<br>(0.4460) | -0.188<br>(0.1534) | -0.123<br>(0.3538) | -0.053<br>(0.6923) | 0.166<br>(0.2081) | 0.046<br>(0.7298) | -0.112<br>(0.3980) |
| WDR78_circ<br>_0006677 | 0.547<br>(0.4596) | -0.086<br>(0.3541) | -0.252<br>(0.0545) | -0.074<br>(0.5794) | -0.037<br>(0.7812) | 0.071<br>(0.5919) | 0.044<br>(0.7407) | 0.001<br>(0.9918) |
| HAT1_circ<br>_0008032 | 4.783<br>(0.0287) | -0.039<br>(0.6710) | -0.098<br>(0.4550) | -0.131<br>(0.3176) | 0.018<br>(0.8914) | 0.100<br>(0.4488) | 0.138<br>(0.2922) | 0.030<br>(0.8196) |
| ATP6V0A1_circ_0043837 | 0.010<br>(0.9208) | 0.048<br>(0.6025) | -0.111<br>(0.3978) | 0.095<br>(0.4719) | -0.001<br>(0.9958) | -0.008<br>(0.9523) | -0.072<br>(0.5843) | 0.070<br>(0.5971) |
| REPS1_circ_0004368 | 0.363<br>(0.5467) | 0.012<br>(0.8933) | 0.108<br>(0.4099) | -0.206<br>(0.1135) | -0.009<br>(0.9453) | -0.148<br>(0.2587) | -0.023<br>(0.8642) | -0.054<br>(0.6831) |
| KIDINS220_circ_0005315 | 4.019<br>(0.0450) | 0.010<br>(0.9103) | -0.093<br>(0.4845) | -0.076<br>(0.5681) | 0.058<br>(0.6643) | 0.227<br>(0.0833) | 0.036<br>(0.7864) | 0.020<br>(0.8785) |
| TMEM138_circ_0002058 | 0.020<br>(0.8864) | 0.032<br>(0.7304) | -0.008<br>(0.9521) | -0.030<br>(0.8227) | 0.014<br>(0.9144) | -0.043<br>(0.7471) | 0.068<br>(0.6034) | 0.030<br>(0.8221) |
| MAPK9_circ_0001566 | 1.954<br>(0.1621) | 0.044<br>(0.6357) | -0.119<br>(0.3663) | 0.063<br>(0.6301) | 0.062<br>(0.6389) | 0.202<br>(0.1208) | 0.057<br>(0.6662) | -0.010<br>(0.9406) |
| KIAA1841_circ_0007793 | 0.034<br>(0.8546) | -0.014<br>(0.8824) | -0.179<br>(0.1715) | -0.027<br>(0.8372) | 0.089<br>(0.4983) | 0.025<br>(0.8473) | 0.050<br>(0.7058) | 0.244<br>(0.0601) |
| TMCC1_circ_0001340 | 0.053<br>(0.8185) | -0.006<br>(0.9453) | -0.104<br>(0.4335) | -0.049<br>(0.7133) | 0.026<br>(0.8473) | 0.093<br>(0.4816) | 0.045<br>(0.7331) | 0.049<br>(0.7141) |
| AGTPBP1_circ_0007162 | 0.068<br>(0.7941) | 0.033<br>(0.7198) | -0.047<br>(0.7215) | -0.076<br>(0.5616) | 0.168<br>(0.1995) | 0.156<br>(0.2330) | 0.110<br>(0.4023) | 0.264<br>(0.0413) |
| PSEN1_circ_0003848 | 0.148<br>(0.7001) | 0.002<br>(0.9865) | -0.263<br>(0.0424) | 0.012<br>(0.9249) | 0.077<br>(0.5594) | 0.160<br>(0.2218) | 0.010<br>(0.9394) | 0.244<br>(0.0606) |
| ZNF292_circ_0004058 | 0.121<br>(0.7283) | -0.020<br>(0.8305) | -0.089<br>(0.5008) | -0.232<br>(0.0770) | -0.022<br>(0.8662) | 0.077<br>(0.5620) | -0.038<br>(0.7733) | -0.074<br>(0.5774) |
| FAM120A_circ_0001875 | 0.047<br>(0.8279) | -0.082<br>(0.3747) | -0.237<br>(0.0687) | -0.145<br>(0.2675) | -0.070<br>(0.5942) | 0.129<br>(0.3253) | -0.041<br>(0.7551) | -0.009<br>(0.9473) |
| FUT8_circ_0003028 | 0.493<br>(0.4826) | -0.074<br>(0.4238) | -0.214<br>(0.1004) | 0.029<br>(0.8233) | -0.040<br>(0.7600) | 0.072<br>(0.5852) | -0.094<br>(0.4761) | -0.114<br>(0.3875) |
| KDM4C_circ_0001839 | 0.030<br>(0.8619) | 0.070<br>(0.4462) | 0.122<br>(0.3525) | -0.183<br>(0.1627) | -0.177<br>(0.1753) | 0.069<br>(0.6028) | -0.153<br>(0.2436) | -0.121<br>(0.3555) |
| HTT_circ_0001392 | 1.317<br>(0.2512) | -0.092<br>(0.3232) | -0.045<br>(0.7327) | -0.101<br>(0.4470) | 0.078<br>(0.5564) | 0.118<br>(0.3754) | 0.054<br>(0.6859) | -0.006<br>(0.9664) |
| MED12L_circ_0067735 | 1.103<br>(0.2937) | -0.137<br>(0.1368) | -0.415<br>(0.0010) | 0.221<br>(0.0893) | 0.066<br>(0.6178) | 0.050<br>(0.7037) | 0.008<br>(0.9512) | 0.246<br>(0.0580) |
| ZFAND6_circ_0000643 | 4.280<br>(0.0386) | -0.053<br>(0.5659) | -0.138<br>(0.2939) | -0.148<br>(0.2585) | -0.046<br>(0.7246) | 0.098<br>(0.4573) | -0.029<br>(0.8284) | -0.108<br>(0.4101) |
| SLAIN2_circ_0126525 | 0.623<br>(0.4301) | 0.015<br>(0.8669) | -0.060<br>(0.6462) | -0.135<br>(0.3030) | -0.100<br>(0.4489) | 0.096<br>(0.4651) | -0.043<br>(0.7458) | -0.039<br>(0.7653) |
| PHC3_circ_0001359 | 0.418<br>(0.5182) | -0.003<br>(0.9738) | -0.064<br>(0.6282) | -0.217<br>(0.0951) | -0.068<br>(0.6062) | 0.139<br>(0.2897) | -0.037<br>(0.7778) | -0.077<br>(0.5598) |

**Significance Supplemental Table 5.** KEGG and GOslim categories that are mostly deregulated in idiopathic PD. Gene union of the targets of the five miRNAs (miR-516b-5p, miR-526b-5p, miR-578, miR-659-3p, miR-1197) sequestered by half or more of the deregulated PD circRNAs in PD versus (A) KEGG and (B) GOslim categories created by the DIANA-miRPath v3.0 interface using default values (*p*-value threshold 0.05, microT-CDS threshold 0.8).

**A**
| KEGG pathway | *P* value | # genes | # miRNAs |
| --- | --- | --- | --- |
| Thyroid hormone signaling pathway | 0.001458 | 24 | 5 |
| Cell cycle | 0.009263 | 25 | 5 |
| Regulation of actin cytoskeleton | 0.014546 | 42 | 5 |
| Rap1 signaling pathway | 0.01546 | 35 | 5 |
| Phosphatidylinositol signaling system | 0.015883 | 16 | 4 |
| MAPK signaling pathway | 0.015883 | 46 | 5 |
| FoxO signaling pathway | 0.015883 | 26 | 5 |

**B**
| GOslim category | *P* value | # genes | # miRNAs |
| --- | --- | --- | --- |
| Organelle | 1.13E-78 | 1143 | 5 |
| Cellular nitrogen compound metabolic process | 6.05E-47 | 587 | 5 |
| Cellular protein modification process | 3.93E-19 | 283 | 5 |
| Nucleoplasm | 2.34E-13 | 159 | 5 |
| Enzyme binding | 2.34E-13 | 168 | 5 |
| Protein complex | 4.38E-11 | 411 | 5 |
| Protein binding transcription factor activity | 6.74E-10 | 70 | 5 |
| Cytoskeletal protein binding | 3.91E-09 | 102 | 5 |
| Cell death | 7.91E-08 | 112 | 5 |
| RNA binding | 6.93E-07 | 207 | 5 |
| Nucleic acid binding transcription factor activity | 7.74E-07 | 112 | 5 |
| Macromolecular complex assembly | 3.06E-05 | 95 | 5 |
| Response to stress | 3.72E-05 | 225 | 5 |
| Membrane organization | 4.89E-05 | 65 | 5 |
| Cytoskeleton organization | 0.000178 | 80 | 5 |

